# Gut Microbiome Structure and Progression in Multiple Sclerosis: A Meta-Analysis Across 14 COHORTS

**DOI:** 10.1101/2025.09.08.25335213

**Authors:** Shauni Doms, John F. Baines, Jerome Hendriks

## Abstract

The gut microbiome is increasingly implicated in multiple sclerosis (MS), yet findings across studies remain inconsistent due to small sample sizes and methodological variability. We performed the largest MS microbiome meta-analysis to date, integrating 14 datasets (n = 1,493; 777 RRMS, 87 PPMS, 66 SPMS, 563 healthy controls) with standardized preprocessing. We analyzed community diversity, taxonomic composition, co-occurrence networks, and enterotype clustering, accounting for technical and demographic confounders. Alpha diversity did not differ significantly between MS subtypes, treatment groups, or controls. However, beta diversity revealed small, but significant differences between MS patients and controls, and among MS subtypes. Differential abundance analysis identified taxa enriched in MS, *Akkermansia,* and *Eisenbergiella*, which were notably associated with progressive disease. A random forest classifier distinguished RRMS from progressive MS with ∼84% accuracy. Co-occurrence networks differed by subtype: RRMS networks were fragmented, SPMS were more cohesive, and PPMS were sparse, but highly modular. Hub taxa composition shifted accordingly. Enterotyping revealed five clusters; RRMS samples were enriched in *Akkermansia*- and *Blautia*-dominated types, while controls showed enrichment in *Faecalibacterium*- and Christensenellaceae-rich clusters. Progressive MS lacked a dominant enterotype. Together, these findings support a “fragmentation hypothesis” in which microbial ecosystems become increasingly unstable with MS progression.

## INTRODUCTION

Multiple sclerosis (MS) is a chronic, immune-mediated neurodegenerative disorder characterized by inflammatory attacks on myelin, and in later stages on axons themselves. MS presents in several clinical subtypes, which are crucial for determining both prognosis and treatment strategies. The most common form, relapsing-remitting MS (RRMS), accounts for about 85% of diagnoses and is defined by episodes of neurological symptoms (relapses) followed by partial or complete recovery (remissions), with no evident progression during remission periods. RRMS often progresses into secondary progressive MS (SPMS), marked by a steady decline in neurological function with no remissions. A smaller subset of patients (∼10– 15%) present with primary progressive MS (PPMS), characterized by continuous worsening from the beginning without distinct relapses. The rarest subtype, progressive-relapsing MS (PRMS), involves gradual progression from the start, with acute relapses happening intermittently.

Notably, up to two-thirds of people with MS (pwMS) report gastrointestinal symptoms before neurological onset, highlighting a potential role of the gut and its microbiome in disease development. This has led to a surge in microbiome studies in MS over the past decade. Several studies have reported significant differences in gut microbial composition between pwMS and healthy controls. Additionally, several studies suggest that disease-modifying therapies (DMTs) can modulate gut microbiota composition (Castillo-Álvarez et al. 2021; Troci et al. 2022; Moles et al. 2024; Gupta et al. 2025). However, most of these studies are limited by small sample sizes, heterogeneous methodologies, and inconsistent findings.

To overcome these limitations, we leveraged all publicly available 16S rRNA gene sequencing datasets from MS studies, integrating accompanying metadata on MS subtype, treatment status, demographics, and technical variables. By applying a standardized data processing and analysis pipeline, we explicitly controlled for major methodological confounders such as sequencing region and DNA extraction method. With approximately 1,800 samples, this work represents the largest meta-analysis of the MS gut microbiome to date, enabling a robust assessment of microbial shifts associated with MS and its subtypes, and providing new insight into the interplay between disease status, treatment, and technical variation in microbiome profiling.

## RESULTS

### Data overview

In total, we included 13 different publicly available MS microbiome studies and one in-house dataset, resulting in a total of 971 pwMS (778 RRMS, 90 PPMS, 67 SPMS, 2 atypical MS, and 40 unknown) and 767 healthy controls (HC; Suppl. Table 1). After read processing and quality filtering, 563 HC, 777 RRMS, 87 PPMS, and 66 SPMS individuals remain (**Table 1**).

**Table 1:** Summary statistics of the samples included in the meta-analysis.

| Variable | N | Control<br>N = 563<sup>1</sup> | PPMS<br>N = 87<sup>1</sup> | RRMS<br>N = 777<sup>1</sup> | SPMS<br>N = 66<sup>1</sup> |
| --- | --- | --- | --- | --- | --- |
| **Age_years** | 1,236 | 51 (41, 61) | 59 (53, 64) | 44 (35, 53) | 59 (53, 64) |
| Unknown |  | 13 | 11 | 233 | 0 |
| **BMI** | 1,329 | 26.3 (23.6, 29.1) | 23.9 (21.4, 27.9) | 25.0 (21.9, 29.0) | 24.8 (21.9, 27.9) |
| Unknown |  | 13 | 15 | 135 | 1 |
| **Disease_duration_months** | 696 | NA (NA, NA) | 120 (72, 198) | 108 (36, 192) | 324 (204, 420) |
| Unknown |  | 563 | 11 | 223 | 0 |
| **Disease_severity_score_EDSS** | 1,118 | 0.00 (0.00, 0.00) | 5.25 (3.50, 6.00) | 1.50 (0.00, 2.50) | 6.00 (4.00, 6.50) |
| Unknown |  | 14 | 13 | 348 | 0 |
| **Disease_severity_MSSS** | 1,099 | 0.00 (0.00, 0.00) | 6.69 (5.25, 7.94) | 1.84 (0.64, 3.65) | 6.06 (4.51, 7.23) |
| Unknown |  | 14 | 23 | 356 | 1 |

<sup>1</sup> Median (Q1, Q3)

### Microbiome composition across studies

To identify patterns between cases and controls, we combined the microbiome data from all studies at the genus level. We found no significant difference in alpha diversity using Shannon (richness and evenness; **Figure 1**) and Chao1 (species richness; Suppl. Fig. 1) diversity indices between pwMS and healthy controls (**Figure 1**A), between treated and untreated pwMS (**Figure 1**B), between MS types (**Figure 1**C), or between treatment drugs (**Figure 1**D), using a linear mixed model with Project ID and the sequencing region as random effects. In contrast, for beta diversity we observe a significant difference in microbial composition between pwMS and HC (PERMANOVA, p=0.001, 1000 permutations; **Figure 2**A) and the different MS types (PERMANOVA, p=0.003, 1000 permutations; **Figure 2**B).

**Figure 1:**
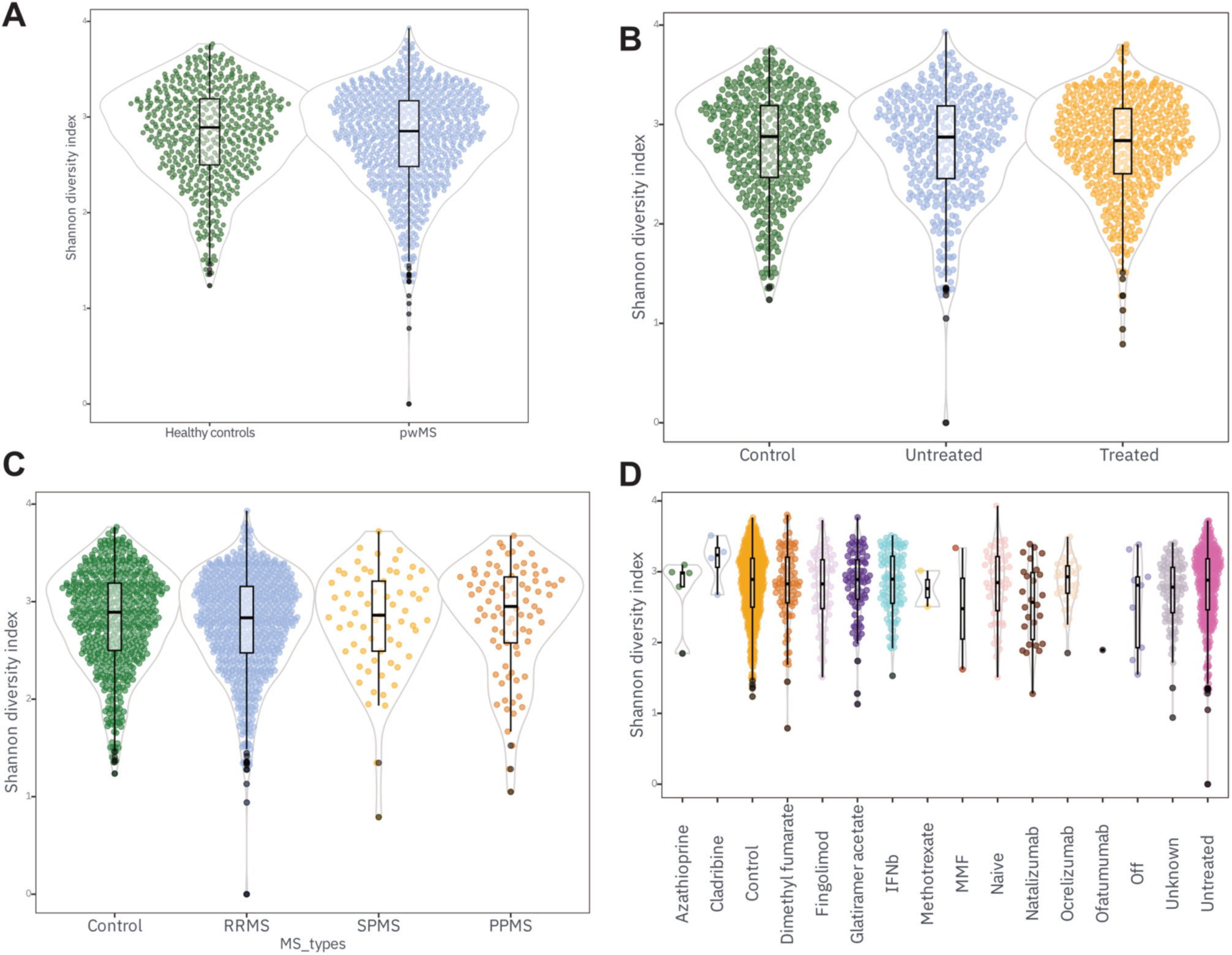
Comparison of alpha diversity across MS disease status, treatment, and subtypes. (A) Alpha diversity between people with MS (pwMS) and healthy controls. (B) Alpha diversity among healthy controls, untreated pwMS, and treated pwMS. (C) Alpha diversity across clinical MS subtypes (CIS, RRMS, SPMS, PPMS). (D) Alpha diversity stratified by disease-modifying treatment. Shannon diversity was used as the metric of alpha diversity. Statistical significance was assessed using linear mixed-effects models, with alpha diversity as the outcome and Project ID included as a random effect to account for cohort-specific variation. The Tukey post hoc test was used to test the pairwise comparisons. None of the comparisons were significant.

**Figure 2:**
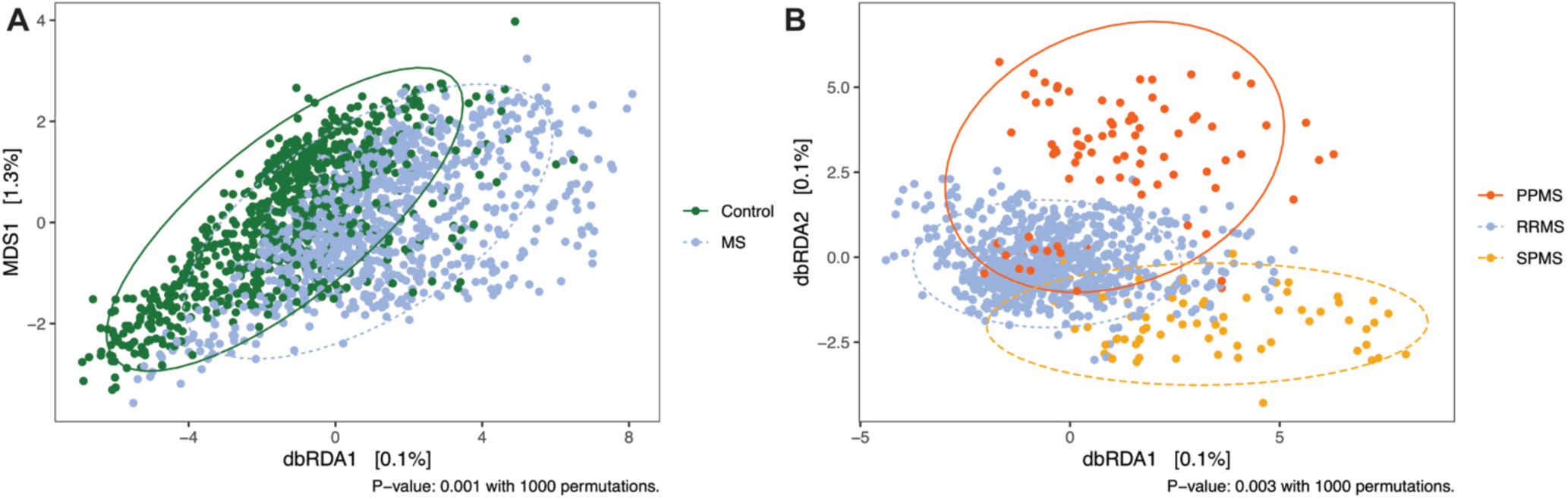
Disease status and MS types explain a small but significant portion of gut microbial variation after accounting for technical and demographic factors. Distance-based redundancy analysis (db-RDA) of Bray-Curtis dissimilarities reveals the effect of MS status (A) or MS types (B) on gut microbiome composition, while conditioning on technical covariates, including project ID and sequencing region. Each point represents a sample, colored by (A) MS status or (B) MS type. Axes represent the constrained variance explained by disease status (A) or MS type (B), with residual variation removed through partial conditioning. Although disease status and MS types account for a modest proportion of total variance, the separation is statistically significant (PERMANOVA, HC vs pwMS: p=0.001; MS types: p=0.003, 1000 permutations), supporting a subtle but consistent disease-associated shift in community structure.

### Differentially abundant taxa

We next identified 34 taxa that were significantly differentially abundant between healthy controls and pwMS. Among these, twelve taxa were increased in pwMS with a fold change of at least 1.5, including *Akkermansia*, *Hungatella*, *Lactobacillus*, *Ruthenibacterium*, *Eisenbergiella*, *Gemella*, *Anaerotruncus*, *Clostridium innocuum* group, and *Frisingicoccus* (**Figure 3**A). Conversely, three taxa were significantly decreased in pwMS (fold change < - 1.5): *Catenibacter*, *Segatella*, and an unclassified genus within the *Prevotellaceae* family (**Figure 3**A). We then stratified the analysis by MS subtype and compared each group to healthy controls. This revealed 28 distinct differentially abundant taxa across the three subtypes (**Figure 3**B): 15 in relapsing-remitting MS (RRMS), 15 in primary progressive MS (PPMS), and 17 in secondary progressive MS (SPMS). Of these, 23 taxa exhibited a fold change greater than ±1.5 (4 in RRMS, 10 in PPMS, and 16 in SPMS). Of particular interest are the genera *Akkermansia* (**Figure 3**C) and *Faecalibacterium*(**Figure 3**D), both of which have been previously associated with numerous human diseases (Pellegrino et al. 2023; Martín et al. 2023). In treated pwMS, only one genus (*CAG-352*) was significantly increased, while two genera (*UCG-002* and *UGC-005*) were decreased (**Figure 3**E). Finally, to assess the impact of disease progression, we combined the PPMS and SPMS groups and identified seven differentially abundant taxa (**Figure 3**F). Four genera were increased in progressive MS (*Akkermansia*, *Eisenbergiella*, *NK4A214 group*, and *Intestinimonas*), while three were decreased: *Blautia*, *Lachnoclostridium*, and *Dialister*.

**Figure 3:**
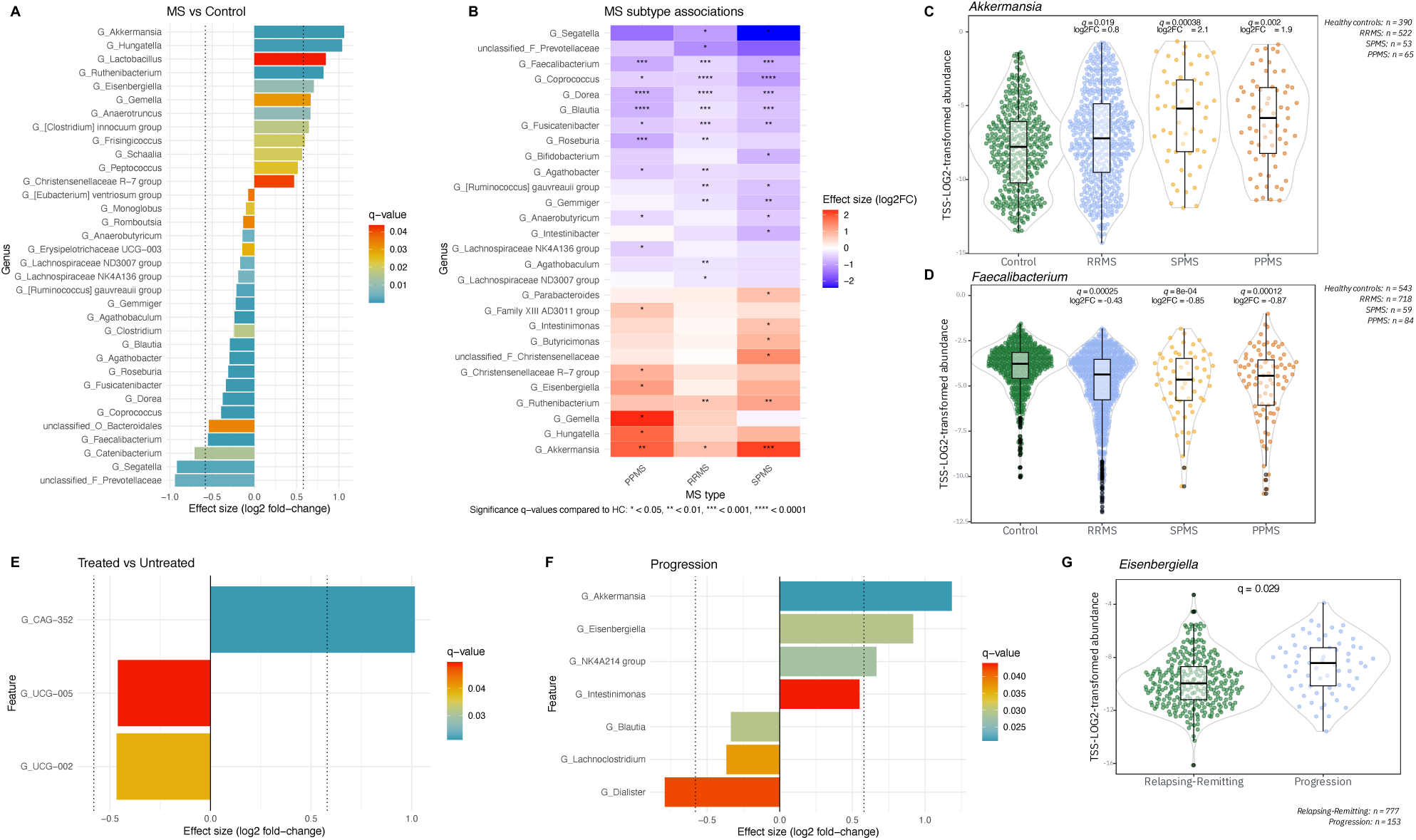
Differentially abundant genera between healthy controls and MS groups. (A) Genera significantly different between people with MS (pwMS) and healthy controls (HC), based on log2 fold change and q-value. (B) Differentially abundant genera per MS subtype (RRMS, PPMS, SPMS) compared to HC. The abundance of Akkermansia (C) or Faecalibacterium (D) in MS subtypes. Q-values and log2FC are relative to healthy controls. (E) Genera that are significantly differentially abundant between treated and untreated pwMS. (F) Genera that are significantly differentially abundant in progressive disease (SPMS and PPMS) compared to RRMS. (G) Abundance of genus Eisenbergiella in progressive vs relapsing-remitting disease. Significance codes: q < 0.05 (*), < 0.01 (**), < 0.001 (***), < 0.0001 (****).

### Prediction of progressive disease by microbiome

We trained a random forest classifier to distinguish between relapsing-remitting MS (RRMS) and progressive MS (SPMS and PPMS) using CLR-transformed genus-level gut microbial profiles. The model achieved an out-of-bag (OOB) error rate of 16.3%, corresponding to a classification accuracy of approximately 83.7%. This indicates that the gut microbiome harbors a predictive signal related to MS progression status. Feature importance analysis revealed that several microbial genera contributed strongly to the disease classification, including UGC-005, *Akkermansia,* and *Intestinimonas* (Suppl. Fig. 1), which were also identified in the differential abundance analysis above. Thus, these taxa may represent important microbiomarkers of disease progression.

### Gut microbial networks differ across MS subtypes

Next we performed pairwise co-occurrence network comparisons of RRMS, SPMS, and PPMS gut microbiomes, which revealed distinct architectures for each MS subtype (**Figure 4A–C**). RRMS networks were moderately modular and weakly clustered, consistent with a more fragmented and sparsely connected microbial structure (Control_vs_RRMS). SPMS, which typically follows RRMS in disease progression, exhibited greater modularity and clustering (Control_vs_SPMS, RRMS_vs_SPMS), suggesting a more cohesive microbial configuration. In contrast, PPMS networks were highly modular, but sparse, with minimal clustering and fewer positive associations (Control_vs_PPMS, RRMS_vs_PPMS, SPMS_vs_PPMS) reflecting a segregated and possibly dysbiotic microbial community.

**Figure 4:**
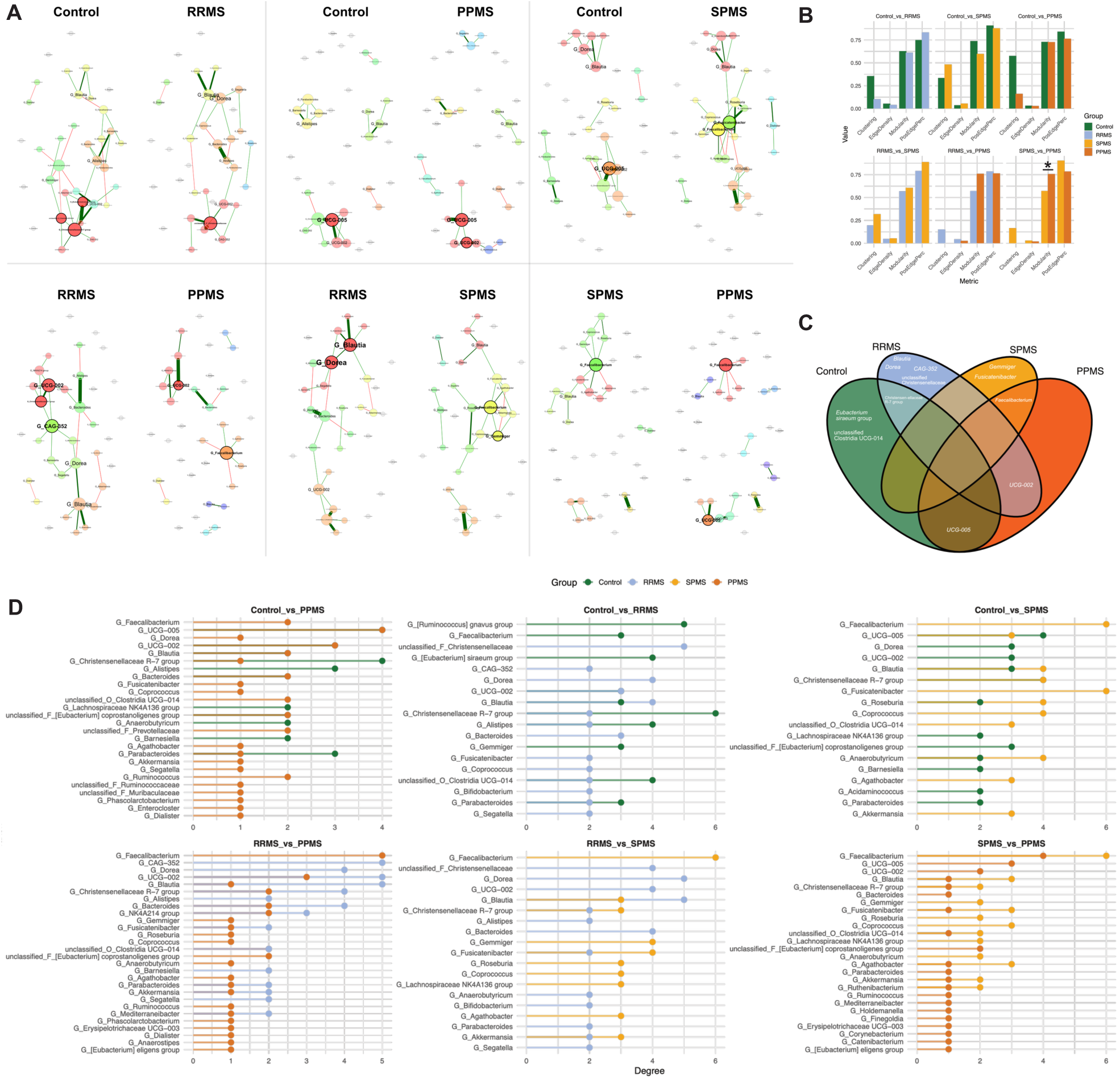
Co-occurrence networks and centrality differences across MS subtypes and controls. (A) Microbial co-occurrence networks were constructed for pairwise comparisons between MS subtypes (RRMS, SPMS, PPMS) and healthy controls using the SPRING method and filtered for the 50 most variable genera. Nodes represent genera, with size proportional to degree centrality and color indicating cluster membership. The layout of the nodes is calculated as a union of both networks to allow for easier comparison. (B) Global network metrics, including clustering coefficient, edge density, modularity, and percentage of positive associations, were calculated for each group. Statistical differences were determined using n=10,000 permutations. * p<0.05 (C) Venn diagram depicting hub taxa in the MS subtype-specific networks. (D) Degree centrality values of key hub taxa across groups.

To statistically assess these differences, we performed permutation-based testing (n = 1000) on global network properties (Suppl. Table X). At the full-network level, only one pairwise comparison, PPMS vs SPMS, showed a significant difference in modularity (p = 0.048). Low ARI p-values suggest significant differences in microbial community clustering between MS subtypes, particularly between RRMS and SPMS. This is further supported by a significantly low Jaccard index of hub taxa for the same comparison (*p* = 0.027), indicating a shift in core microbial players. Despite high GCD values across comparisons, their non-significant p-values suggest that large-scale topological differences, while present, may not be consistent enough to reach statistical significance across samples.

To focus on the most structurally relevant parts of the networks, we repeated our analysis on the largest connected component (LCC). Differences were more pronounced in this core subnetwork. The RRMS vs PPMS comparison showed significantly higher modularity (p = 0.017) and reduced natural connectivity (p = 0.028) in PPMS, indicating a more compartmentalized, yet fragile microbial core. Edge density was also lower in PPMS (p = 0.099), pointing to fewer interactions among central taxa.

Finally, we assessed hub taxa using degree centrality. Hub compositions differed substantially across groups (**Figure 4C**). *Faecalibacterium* had significantly higher centrality in both SPMS and PPMS compared to RRMS (p = 0.098 and 0.044, resp.), while *Gemmiger* was more central in RRMS than in controls (p = 0.007) (**Figure 4D**). *Christensenellaceae R-7* group, a shared hub in controls and RRMS (**Figure 4C**), did not differ significantly between subtypes (p > 0.2). Other genera with subtype-specific centrality shifts included *Alistipes*, more central in PPMS than controls (p = 0.013), and *Dorea* and *CAG-352*, both more central in PPMS than RRMS (p = 0.082 and 0.005, respectively).

### Enterotype enrichment in MS subtypes

To further explore gut microbial community structure, we performed enterotyping using Dirichlet multinomial mixture modeling (Holmes et al. 2012; Costea et al. 2018) and identified five distinct enterotype clusters across all samples. We then examined the distribution of these enterotypes across MS subtypes and controls (**Figure 5A**), as well as the dominant taxa characterizing each enterotype (**Figure 5B**). Enterotype 1 was mostly dominated by

**Figure 5:**
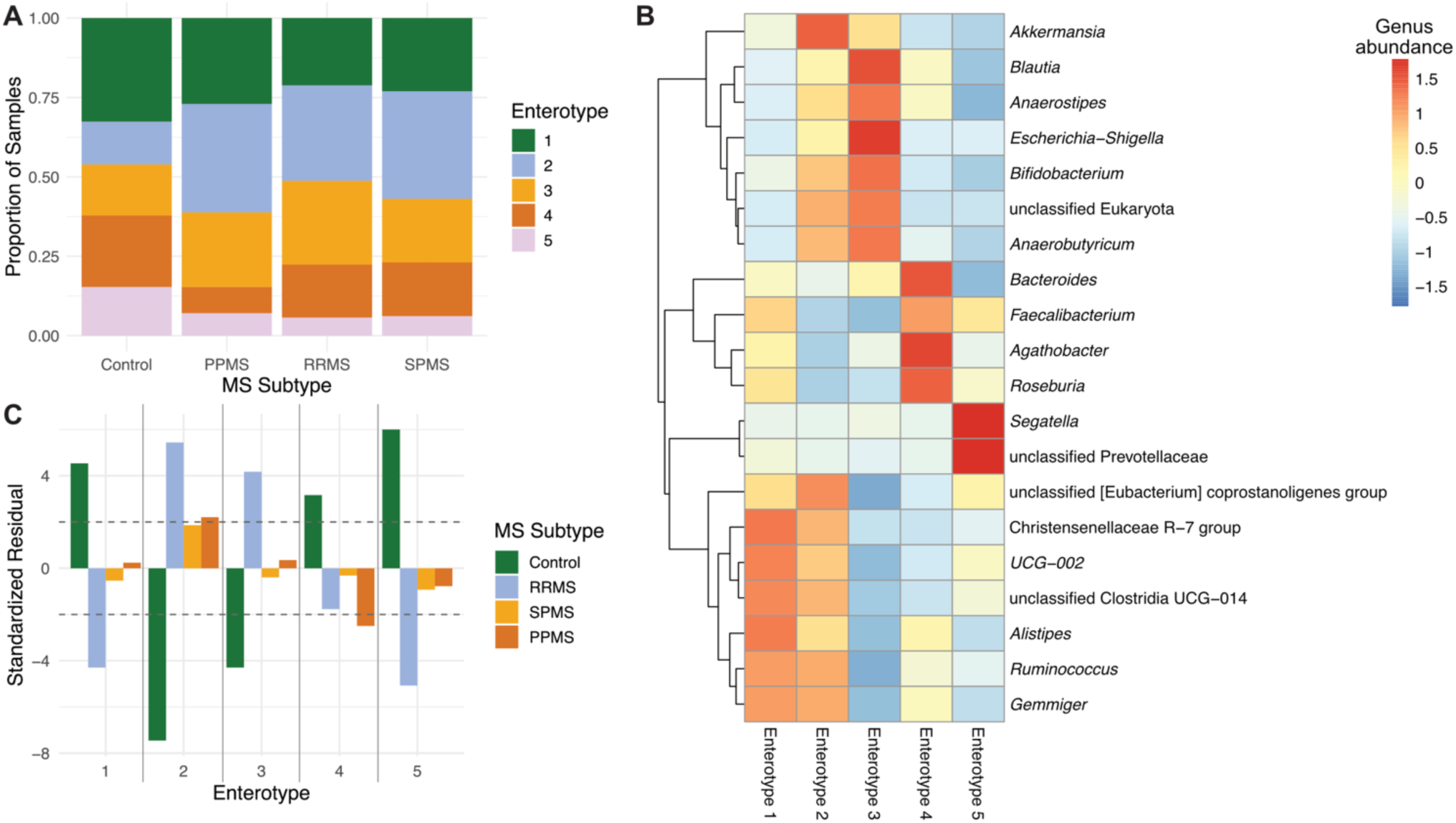
Enterotype classification across MS subtypes. (A) Proportion of enterotypes across MS subtypes. (B) Top 20 variable genera across enterotypes. Genus abundances are depicted as the center-scaled averages in the enterotype. (C) Standardized residuals of the Chi-Square test showing the enrichment of an enterotype in the MS subtype. The dashed horizontal line indicates the threshold for significance (|z|>2).

Christensenellaceae R-7 group, UGC-002, an unclassified genus belonging to Clostridia UCG-014, and *Alistipes*. Enterotype 2 was strongly enriched in *Akkermansia*. Enterotype 3 featured high abundances of *Blautia*, *Anaerostipes*, *Escherichia*-*Shigella*, *Bifidobacterium*, and *Anaerobutyricum*. Enterotype 4 was characterized by *Bacteroides*, *Faecalibacterium*, *Agathobacter*, and *Roseburia*. Finally, enterotype 5 was dominated by *Segatella* and an uncharacterized genus belonging to the Prevotellaceae family.

To statistically assess group differences in enterotype frequency, we calculated standardized residuals from a chi-squared test comparing enterotype distributions across MS subtypes. The results revealed significant enrichment of Enterotypes 2 and 3 in RRMS, while healthy controls were overrepresented in Enterotypes 1, 4, and 5 (**Figure 5C**). Specifically, RRMS samples had strong positive residuals in Enterotype 2 (z = 5.4) and Enterotype 3 (z = 4.2), while controls showed significant depletion in those same clusters (z = –7.5 and –4.3, respectively). In contrast, SPMS and PPMS showed no strong or consistent enterotype enrichment, although PPMS exhibited a mild overrepresentation of Enterotype 2 (z = 2.2).

These findings suggest that gut microbial communities in RRMS cluster into distinct compositional profiles that differ markedly from those in healthy controls. In contrast, progressive MS forms appear to exhibit more diffuse or heterogeneous community structures, aligning with our co-occurrence network findings that indicate greater fragmentation and ecological reorganization in late-stage MS.

## DISCUSSION

In this study, we conducted the largest microbiome meta-analysis of multiple sclerosis (MS) to date, integrating 14 datasets and standardizing preprocessing across nearly 1,800 individuals. This harmonized approach enables statistically robust detection of subtle, yet consistent disease-associated microbiome signals, many of which may have been underpowered or inconsistent in smaller, heterogeneous studies.

We observed no significant differences in alpha diversity (Shannon or Chao1) between people with MS (pwMS) and healthy controls, nor across MS subtypes or treatment groups. This finding aligns with the majority of previous studies reporting no consistent alpha diversity shift in MS (Chen et al. 2016; Jangi et al. 2016; Berer et al. 2017; Cekanaviciute et al. 2017; Zeng et al. 2019; Zhou et al. 2022b; Thirion et al. 2023; Devolder et al. 2023; Montgomery et al. 2024). Although earlier work reported decreased alpha diversity in MS (Miyake et al. 2015; Ní Choileáin et al. 2019), our results reinforce the growing consensus that alpha diversity alone is insufficient to reflect health or disease status (Williams et al. 2024).

In contrast, we identified small but statistically significant differences in beta diversity between pwMS and controls, and among MS subtypes. These results confirm that microbial community composition is altered in MS, albeit with modest effect sizes, suggesting that specific taxa or ecological interactions, rather than global diversity shifts, may be more biologically meaningful.

Our differential abundance analysis identified several genera consistently altered in MS, including *Akkermansia*, *Faecalibacterium*, *Hungatella*, and *Eisenbergiella*. Notably, *Akkermansia* and *Eisenbergiella* were enriched in progressive disease (PPMS and SPMS), supporting the hypothesis that disease progression is associated with distinct microbial signatures, rather than a shared MS microbiome profile. *Akkermansia* is known for its mucin-degrading capacity (Ottman et al. 2017) and immunomodulatory effects (Pellegrino et al. 2023), which may have implications for barrier function (Mo et al. 2024), inflammation (Zhao et al. 2024), and host-microbe signaling (Yaghoubfar et al. 2020; Zhang et al. 2023).

*Eisenbergiella tayi* was recently identified as a likely causal microbe in experimental autoimmune encephalomyelitis (EAE) (Yoon et al. 2025).

Using a random forest classifier, we showed that gut microbial profiles can predict MS subtype, distinguishing RRMS from progressive MS with 84% accuracy. Predictive features included *Akkermansia*, *Intestinimonas*, and UGC-005, indicating that MS progression is accompanied by microbiome shifts with diagnostic potential. While machine learning in microbiome classification is often overfit, our cross-validated performance suggests a real disease signal, although validation in prospective or longitudinal datasets is needed.

To understand microbial community structure, we applied co-occurrence network analysis across MS subtypes. RRMS microbiomes exhibited fragmented and weakly clustered networks, suggesting reduced microbial interaction density in early disease. In contrast, SPMS networks were more cohesive and modular, possibly reflecting reorganization of the microbial ecosystem in response to chronic inflammation, immunomodulatory treatment, or ecological adaptation. PPMS displayed a distinct pattern: high modularity but sparse and disconnected networks, with fewer positive associations—hallmarks of ecological instability and dysbiosis.

These network-level changes suggest that disease progression involves not just compositional shifts, but a reconfiguration of microbial ecosystem architecture. Increased modularity and reduced connectivity may indicate a breakdown in cross-feeding relationships and interdependence among microbes, weakening community resilience. Changes in hub taxa further support this: *Faecalibacterium* became more central in SPMS and PPMS, *Gemmiger* was dominant in RRMS, and Christensenellaceae R-7 group remained stable across subtypes. These may represent dynamic shifts in keystone species whose influence on ecosystem function is critical. The overall trend of fragmentation and segregation, supported by low Jaccard similarity of hub taxa and significant differences in clustering structure (ARI), points toward a loss of ecological redundancy and functional buffering capacity as MS progresses.

Our enterotyping analysis revealed further stratification. RRMS samples were enriched in Enterotypes 2 and 3, dominated by *Akkermansia*, *Blautia*, and *Escherichia*-*Shigella*, while controls were overrepresented in Enterotypes 1, 4, and 5, which included *Faecalibacterium*, *Alistipes*, and *Roseburia*, taxa commonly associated with intestinal health and anti-inflammatory potential (Parker et al. 2020; Nie et al. 2021; Martín et al. 2023). These patterns suggest that RRMS is associated with distinct compositional clusters, while SPMS and PPMS show no clear enterotype enrichment, pointing toward increased community heterogeneity or collapse of ecological structure in later disease stages. This aligns with our network findings, supporting a “fragmentation hypothesis”, in which the microbial ecosystem loses cohesion and resilience as MS progresses.

Our findings highlight the potential of gut microbial features, such as subtype-specific taxa, interaction networks, and enterotype profiles, as informative markers of disease stage and progression in MS. The ability to classify relapsing versus progressive MS based on microbiome data underscores the relevance of these ecological changes for stratifying patients or potentially guiding therapeutic monitoring. However, whether these microbial shifts are causal, compensatory, or simply reflective of host disease state remains to be determined.

Despite harmonized processing and inclusion of technical covariates (e.g., sequencing region, DNA extraction kit), residual batch effects and cohort heterogeneity remain limitations. Future studies with standardized protocols or longitudinal within-cohort designs are essential to validate these patterns. Likewise, while we avoided functional prediction tools due to current limitations in inference accuracy, integrating metagenomics, metatranscriptomics, or metabolomics would help clarify the functional implications of observed taxonomic and network shifts.

Ultimately, this study supports a “fragmentation hypothesis” for the MS microbiome: as the disease progresses, gut microbial communities lose compositional and structural cohesion, becoming increasingly modular, sparse, and subtype-specific. These changes may reflect and potentially influence immune dysregulation and neurodegeneration in MS. Together, our results underscore the importance of moving beyond diversity metrics and focusing on community-level interactions and ecological resilience to understand the role of the microbiome in MS.

## METHODS

### Data collection

We included all publicly available 16S rRNA gene sequences (SRA and EBI archives) from MS studies and one in-house unpublished study. This resulted in 15 available studies (Chen et al. 2016; Jangi et al. 2016; Cekanaviciute et al. 2017; Zeng et al. 2019; Ní Choileáin et al. 2019; Kozhieva et al. 2019; Takewaki et al. 2020; Cantoni et al. 2022; Troci et al. 2022; Yadav et al. 2022; Zhou et al. 2022a; Elsayed et al. 2023; Boussamet et al. 2024; Montgomery et al. 2024; Schwerdtfeger et al. 2025). We further selected only reads sequenced on the Illumina platform with sufficient quality and metadata.

### Data processing

Available metadata was merged across all studies. Reads were processed per study in the following DADA2 (v 1.36.0) pipeline (Callahan 2016; Callahan et al. 2016b, a). In short, raw sequences were trimmed and quality filtered with a maximum of two ‘expected errors’ allowed per read, paired sequences were merged, and amplicon sequence variants (ASVs) were inferred. The resulting ASV sequence tables were merged across studies and chimeras removed. We classified the taxonomy using Silva v138.2 to classify until the species level (Callahan 2024).

### 16S rRNA gene analysis

ASVs were agglomerated to the genus level to be comparable across studies and sequencing regions. Samples with low read depth (less than 10,000 reads) and rare taxa (present in less than 10% of samples) were removed. We further filtered out only feces samples and samples from unique individuals (removed longitudinal sampling). We used the phyloseq R package (v 1.52.0) to estimate alpha diversity using the Shannon and Chao1 indices, and beta diversity using the Bray-Curtis distance (McMurdie and Holmes 2013).

#### Alpha diversity

Reads were rarefied to 10,000 reads to calculate alpha diversity. Differences in alpha diversity were tested using a linear mixed model (lme4 R package, v 1.1.37) with the alpha diversity measure as outcome, MS subtypes (or MS treatment status or MS treatment drugs) as fixed effect, and the studies and sequencing regions as random effects. Post hoc pairwise comparison was performed using the emmeans R package (v 1.11.1) with the Tukey adjustment.

#### Beta diversity

Beta diversity analysis was performed on the relative abundance of the reads. A PERMANOVA (adonis2; vegan R package, v 2.7.1) was performed to test the difference in beta diversity according to MS status (MS vs Control), including the sequencing regions and studies. A distance-based redundancy analysis was performed, partialling out the effect of the different studies and sequencing regions, to get the MS status (or MS subtype) effect on beta diversity. Significance was determined using 1,000 permutations.

#### Differential abundance

Differentially abundant taxa were calculated using the Maaslin3 for the MS status, MS subtypes, MS treatment, MS treatment drugs, MS severity scores (EDSS, MSSS), and MS progression (Relapsing-Remitting vs. Progression). Studies and sequencing regions were added as random effects.

#### Random forest

We trained a random forest classifier to distinguish between relapsing-remitting and progressive MS using center-log-ratio transformed genus-level gut microbial profiles using the randomForest R package (v 4.7.1.2, ntree=500).

#### Co-occurrence networks

We used the NetCoMi R package (v 1.2.0) to calculate spring networks (SPRING R package, v 1.0.4) for each of the possible comparisons between the control and the subtypes (n=6). To control for sample size effects, we subsampled each MS subtype group to the same number of individuals (n = 66). This ensured comparability of network topologies across groups. The networks were then compared using the netCompare function and tested for significant differences using 1,000 permutations.

#### Enterotype analysis

We applied Dirichlet multinomial mixture (DMM) modeling to identify discrete gut microbiome community types (enterotypes) across MS subtypes and healthy controls. We used the rarefied (n=10,000) genus-level abundance matrix, and we filtered out low-prevalence taxa present in fewer than 10% of samples. Next, we fitted models with 1 to 5 clusters. The best-fitting model was selected based on the minimum Laplace approximation criterion. To characterize the defining genera of each enterotype, we aggregated genus-level abundance across samples by enterotype and visualized the 20 most variable genera using z-score-scaled heatmaps. We assessed whether enterotype distributions differed significantly between MS subtypes and controls using a chi-squared test. Post hoc analysis was performed on standardized residuals to identify group-specific over- or underrepresentation of enterotypes.

## Data Availability

All data produced in the present study are available upon reasonable request to the authors

## SUPPLEMENTARY

**Suppl. Fig. 1:**
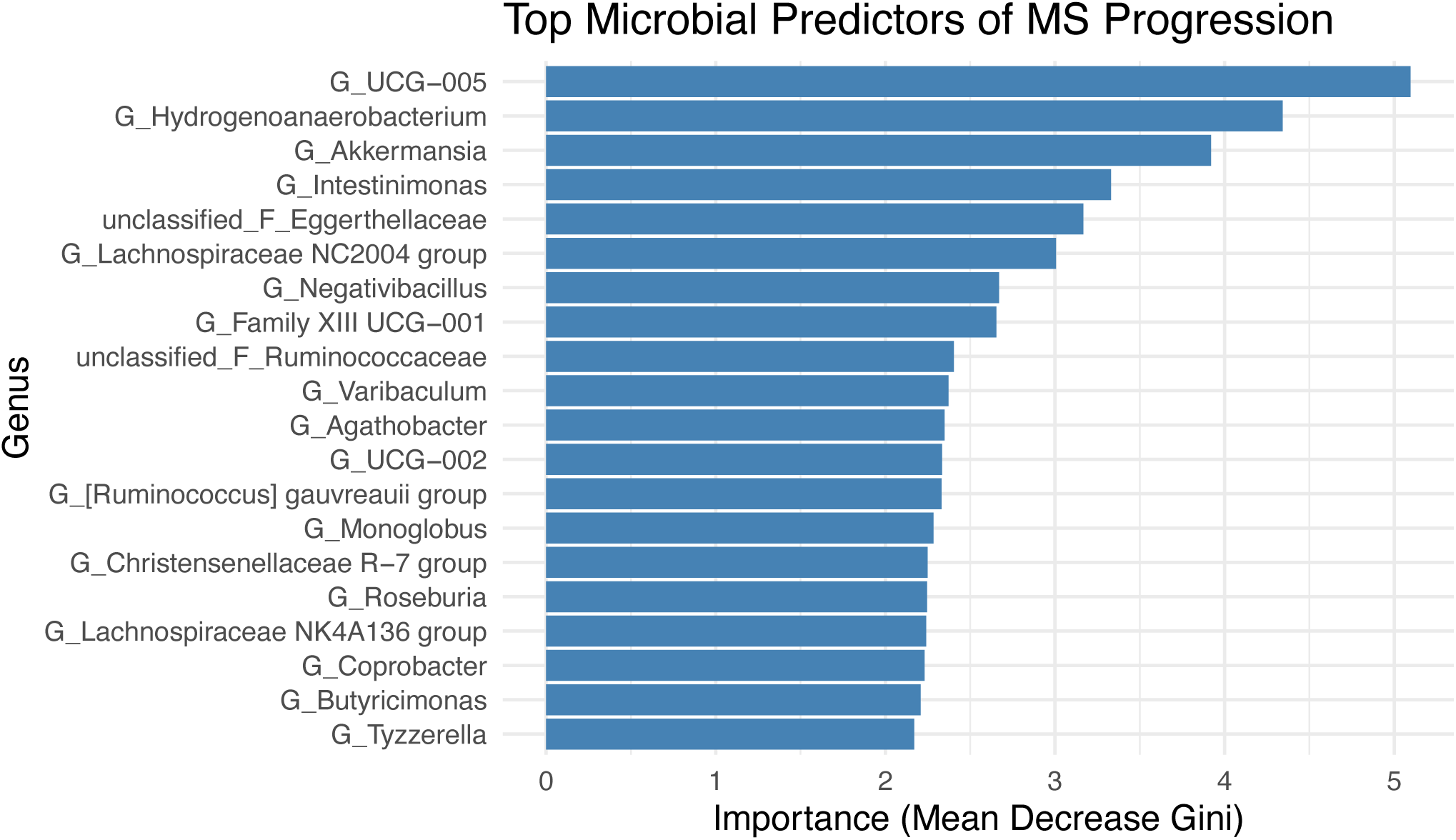
Top 20 microbial predictors of MS progression identified by random forest. The model was trained to distinguish relapsing-remitting MS (RRMS) from progressive MS (SPMS and PPMS) using CLR-transformed genus-level abundances. Bars indicate the Mean Decrease in Gini index, representing each genus’s contribution to classification accuracy. The model achieved an out-of-bag error rate of 16.3%.

